# Excess deaths associated with the Iranian COVID-19 epidemic: a province-level analysis

**DOI:** 10.1101/2020.12.07.20245621

**Authors:** Mahan Ghafari, Alireza Kadivar, Aris Katzourakis

**Author notes:** Corresponding author. (MG); (AK).

## Abstract

**Background:** The number of publicly reported deaths from COVID-19 may underestimate the true death toll from the epidemic as they rely on provisional data that are often incomplete or omit undocumented deaths from COVID-19. In addition, these reports may be subject to significant under-reporting due to a limited testing capacity of a country to identify suspect cases. This study estimated the number of seasonal excess deaths attributable to the COVID-19 epidemic in 31 provinces of Iran.

**Methods:** We gathered the nationwide and provincial time series of the seasonal all-cause mortality data from spring 2015 to summer 2020 (21 March 2015 to 21 September 2020), in accordance with the Solar Hijri (SH) calendar, from the National Organization for Civil Registration (NOCR). We estimated the expected number of seasonal deaths for each province using a piecewise linear regression model which we established based on the mortality figures for the previous years and considered any significant deviations from the expectation during winter, spring, and summer of 2020 to be directly associated with COVID-19.

**Results:** Our analysis shows that from the start of winter to the end of summer (from 22 December 2019 to 21 September 2020), there were a total of 58.9K (95%CI: 46.9K - 69.5K) excess deaths across all 31 provinces with 27% (95%CI: 20% - 34%) estimated nationwide exposure to SARS-CoV-2. In particular, 2 provinces in the central and northern Iran, namely Qom and Golestan, had the highest level of exposure with 57% (95%CI: 44% - 69%) and 56% (95%CI: 44% - 69%), respectively, while another 27 provinces had significant levels of excess mortality in at least one season with >20% population-level exposure to the virus. We also detected unexpectedly high levels of excess mortality during fall 2019 (from 23 September to 21 December 2019) across 18 provinces. Our findings suggest that this spike cannot be a result of an early cryptic transmission of COVID-19 across the country and is also inconsistent with the molecular phylogenetics estimates for the start of the pandemic and its arrival to Iran. However, in the absence of appropriate surveillance data for detecting severe acute respiratory infections we were unable to make a determination as to what caused the spike in fall 2019.

**Conflict of Interest:** None.

## Main

### Historical trends of all-cause mortality in Iran

Information on all-cause mortality in Iran and its evolution over time provides a major source for evaluating the burden of the COVID-19 epidemic across the country. In order to estimate the number of deaths in excess of previous years, i.e. the mortality attributable to a public health crisis, we can compare the total number of registered deaths during the time of the crisis and compare them to some ‘background’ level which is measured during periods with no major crisis [1]. Like many other developing countries, Iran has incomplete records of mortality information. A recent report in 2015 by the United Nations Statistical Division shows that the estimated coverage of registered births and deaths in Iran is 97% and 92%, respectively (coverage information was obtained from country representative(s) either from the National Statistical Office or the Civil Registration Authority attending one of the Nations Statistics Division workshops) [2]. NOCR is responsible for the registration of births, marriages, divorces, and deaths across the country. It initially updates the number of registered (crude) deaths at the end of every season for all the 31 provinces in Iran (according to the Solar Hijri calendar year) [3]. More detailed data on the number of registered deaths per month will then follow at the end of each calendar year (on the March equinox) along with an annual report from the Ministry of Health and Medical Education (MoHME) which includes an aggregate record of causes of deaths (based on ICD10) according to individuals age, sex, and place of residence [4].

Upon examining the historical trends of all-cause mortality in Iran from the 1960s until present, we find significant levels of variations over time (see **Figure 1**). In particular, there is a spike in 1995 with more than 2.7 million registered deaths in just one year (a record-high with almost 15 times higher death toll than previous years). This was part of an ‘emergency operation’ to record significant levels of under-counting from previous years, such as the casualties of the Iran-Iraq war in the 1980s, and redesign NOCR’s data collection strategy [4]. Between 1966 and 1995, the mortality data based on cemetery records were collected only in a sample of 24 cities. In 1995, the system was redesigned to cover the entire country. Since then, the average number of registered deaths has roughly doubled with approximately 389 thousand deaths compared to 175 thousand from previous years. However, even after the completion of the emergency operation (from 1994 to 1999), the annual numbers continue to fluctuate significantly over time with amplitude variations as large as 100 thousand deaths between consecutive years. Such variations are so high that even some of the major natural catastrophes in Iran such as the Manjil-Rudbar earthquake with more than 40 thousand reported deaths in 1990 or the Bam earthquake with more than 50 thousand deaths in 2003 would not have a detectable footprint in the annual all-cause mortality data. Previous works have shown despite the progress since 1995 in reporting the true number of registered deaths, such variations may persist due to substantial delays in registration and recording of deaths [5]. Examining the underlying cause(s) of these large variations is beyond the scope of this study, but we note that this is an area that requires further in-depth investigation.

**Figure 1:**
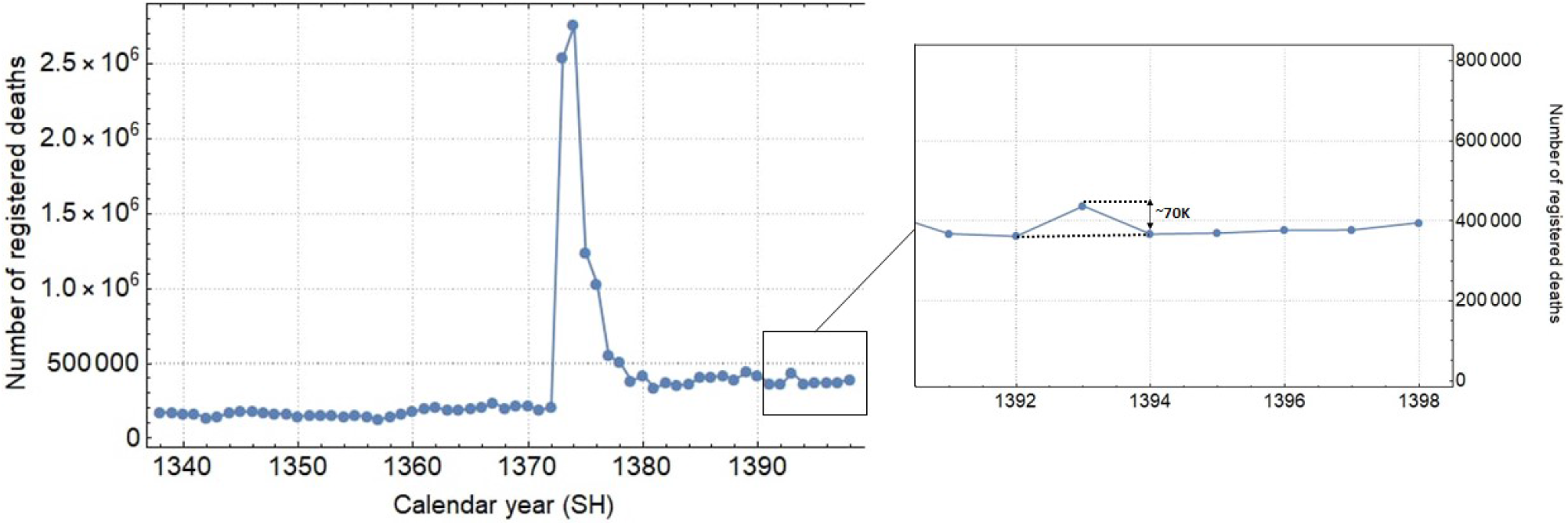
Annual number of registered deaths in Iran over the last 70 years (data from NOCR). The inset shows the variations in registered deaths during the more recent years which seems to have stabilised with the exception of year 1393 SH with approximately 70 thousand deaths above baseline.

MoHME never released provincial data on the number of confirmed COVID-19 deaths and only announced the number of cases for the first 33 days since the start of the epidemic (from 19 February to 22 March 2020, with the exception of 2 and 3 March). Thus, excess mortality is the only proxy to assess the geographic spread of the epidemic across the country. Similar works have also been carried out to monitor excess mortality in Africa during a measles outbreak [6]. Despite significant fluctuations in NOCR’s data over the years, they seem to have remained stable from 1394 SH onward (see **Figure 1**). Therefore, we only incorporate the data from the beginning of spring 1394 SH (21 March 2015) onward to estimate excess mortality rates (see **Methods** section).

### Did the COVID-19 epidemic in Iran start during fall 2019?

Upon examining the seasonal trends in mortality during fall in our earlier reports [7,8], we noticed, for the first time, that there was a spike across the country in 1398 SH (from 23 September to 21 December 2019) compared to previous years. This was particularly unexpected as it occurred before the start of the COVID-19 pandemic in December [9,10] and its emergence in Iran during January [11] which led to some speculations about the cryptic transmission of the virus across the country in fall 2019.

Our analysis shows an 8% rise in nationwide mortality rate during fall with a total of 6.04K (95%CI: 3.48K - 8.60K) extra recorded deaths in 18 provinces -- This is also aligned with estimates from other studies [12]. For these extra deaths in fall to be associated with COVID-19, we would expect, due to the absence of any non-pharmaceutical interventions at the time, that similar (or higher) levels of excess mortality to be detected in the same provinces during winter. However, we find only 5 provinces with significantly higher levels of mortality in winter 1398 SH (22 December 2019 to 19 March 2020) (see **Table 1**). These provinces include Qom, Gilan (Guilan), Golestan, Mazandaran, and Qazvin which were reportedly hit the hardest by an early wave of the COVID-19 epidemic with a total of 3.48K (95%CI: 3.17K - 3.78K) combined extra deaths [13]. This observation rejects any possible association between the excess mortality in fall and significant levels of undetected COVID-19-related deaths across the country.

**Table 1:** Seasonal trend of provinces with (+ sign) or without (- sign) significant levels of excess mortality from fall 1398 SH to summer 1399 SH.

| Provinces | Fall 98 | Winter 98 | Spring 99 | Summer 99 |
| --- | --- | --- | --- | --- |
| East Azerbaijan | + | - | + | + |
| West Azerbaijan | + | - | + | + |
| Ardabil | - | - | + | + |
| Isfahan | + | - | + | + |
| Alborz | - | - | + | + |
| Ilam | + | - | + | + |
| Bushehr | - | - | - | + |
| Tehran | + | - | + | + |
| South Khorasan | - | - | + | - |
| Razavi Khorasan | - | - | + | + |
| North Khorasan | + | - | + | + |
| Khuzestan | - | - | + | + |
| Zanjan | + | - | + | + |
| Semnan | + | - | + | + |
| Sistan and Baluchistan | - | - | - | + |
| Fars | + | - | - | + |
| Qazvin | + | + | + | + |
| Qom | + | + | + | + |
| Kurdistan | + | - | + | + |
| Kerman | - | - | + | + |
| Kermanshah | - | - | + | + |
| Kohgiluyeh and Boyer-Ahmad | - | - | + | + |
| Lorestan | + | - | + | + |
| Mazandaran | + | + | + | + |
| Markazi | + | - | + | + |
| Hormozgan | - | - | + | + |
| Hamedan | - | - | + | + |
| Yazd | + | - | + | + |
| Chaharmahal and Bakhtiari* | - | - | + | - |
| Golestan | + | + | + | + |
| Gilan (Guilan) | + | + | + | + |
| Nationwide | + | - | + | + |

\*We omitted the number of registered deaths for summer 1394 SH out of the analysis for this province as it showed a record-high number with nearly three times more deaths compared to any of the following years.

In search of possible causes of the excess deaths during fall, we investigated the WHO annual influenza situation reports for the Eastern Mediterranean Region in the past two years and found a 20% increase in the number of positive flu cases during fall 1398 SH (with the co-circulation of both H1N1 and flu type B in Iran) [7]. However, in the absence of surveillance data for influenza-like illnesses or severe acute respiratory infections in Iran it is not possible to assess the burden of the flu epidemic across the country.

### The geography of COVID-19 spread in Iran

While the mean nationwide excess mortality was only at approximately 2% during winter, it rose to 21% during spring 1399 SH (from 20 March to 20 June 2020) with a total of 18.36K (95%CI: 13.51K - 23.21K) excess deaths in 28 provinces and 39% during summer 1399 SH (from 21 June to 21 September 2020) with a total of 36.28K (95%CI: 30.26K - 42.30K) excess deaths in 30 provinces (see **Figure 2**).

**Figure 2:**
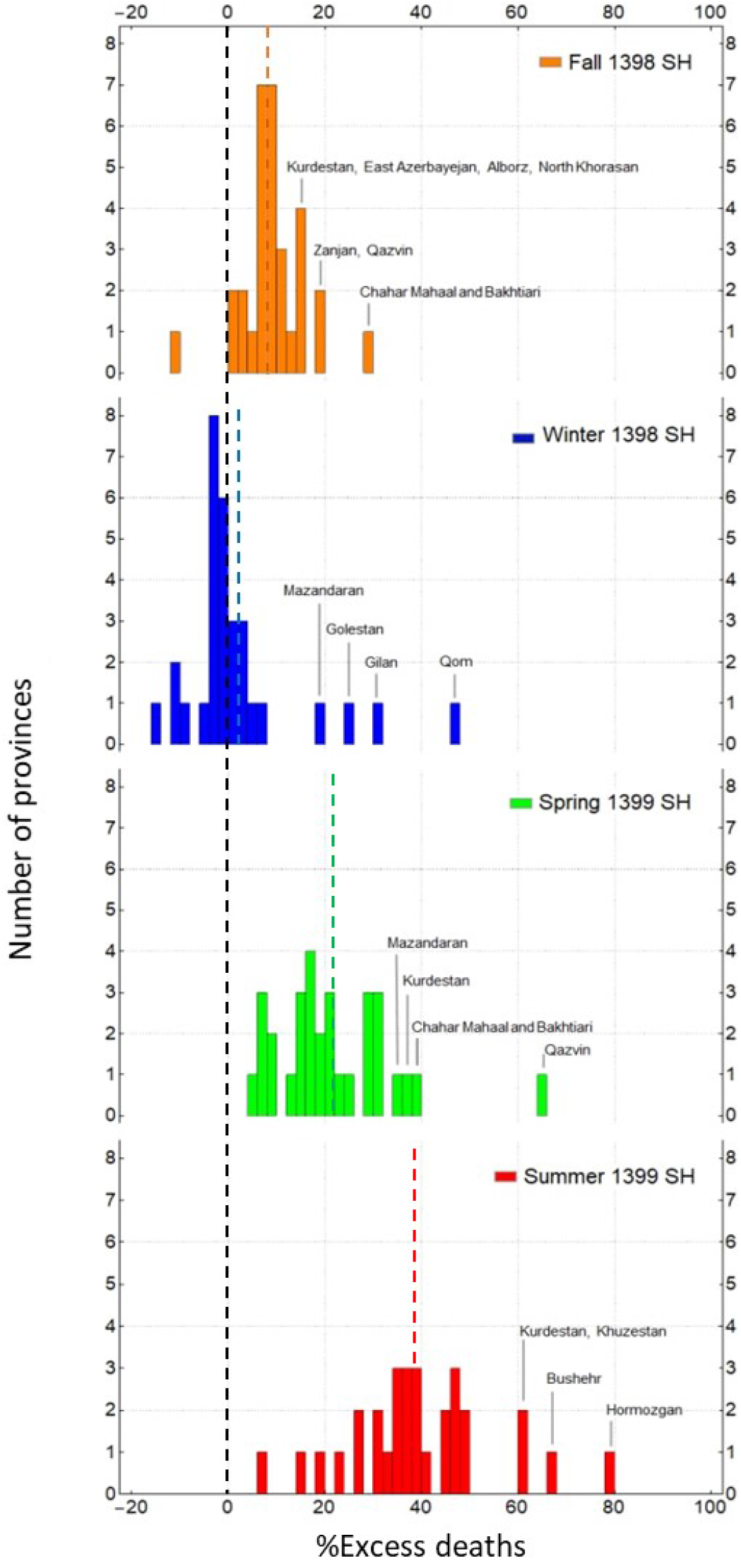
Percentage of excess deaths (compared to the background level) in fall 1398 SH (from 23 September to 21 December 2019), winter 1398 SH (from 22 December to 19 March 2020), spring 1399 SH (from 20 March to 20 June 2020), and summer 1399 SH (from 21 June to 21 September 2020). Some of the provinces with the highest level of excess mortality in each season are highlighted. The vertical dashed lines show the mean percentage of excess mortality in fall (orange), winter (blue), spring (green), and summer (red).

We also find that the 5 provinces in central north part of Iran with an early outbreak of COVID-19 in winter (Qom, Gilan, Golestan, Mazandaran, and Qazvin) continued to have significantly high levels of mortality during spring and summer (see **Figure 3 a**) while 3 provinces in the south (Fars, Bushehr, and Sistan-Baluchistan) only showed a rise in mortality during summer (see **Figure 3 b**). The remaining provinces had significant levels of excess mortality in spring followed by a higher increase during summer (with the exception of South Khorasan which only showed significant excess deaths in spring) which indicates that a sustained and uncontrolled transmission of the virus had been going on for at least six months in most provinces (see **Figure 3 c**). We also note that the level of excess mortality in Gilan continued to decrease after its initial peak during winter while Qom seemed to have increased again in summer after a temporary drop during spring.

**Figure 3:**
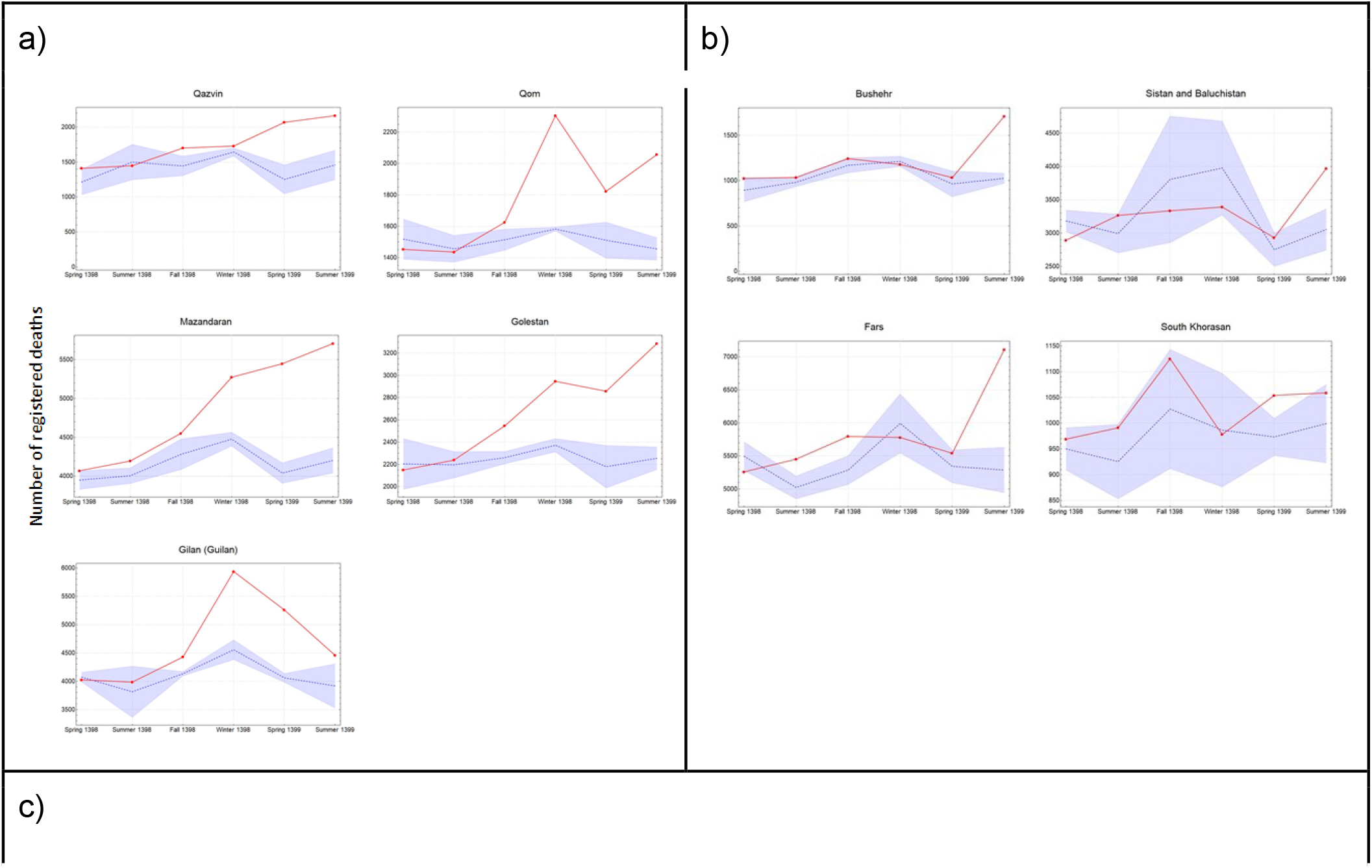
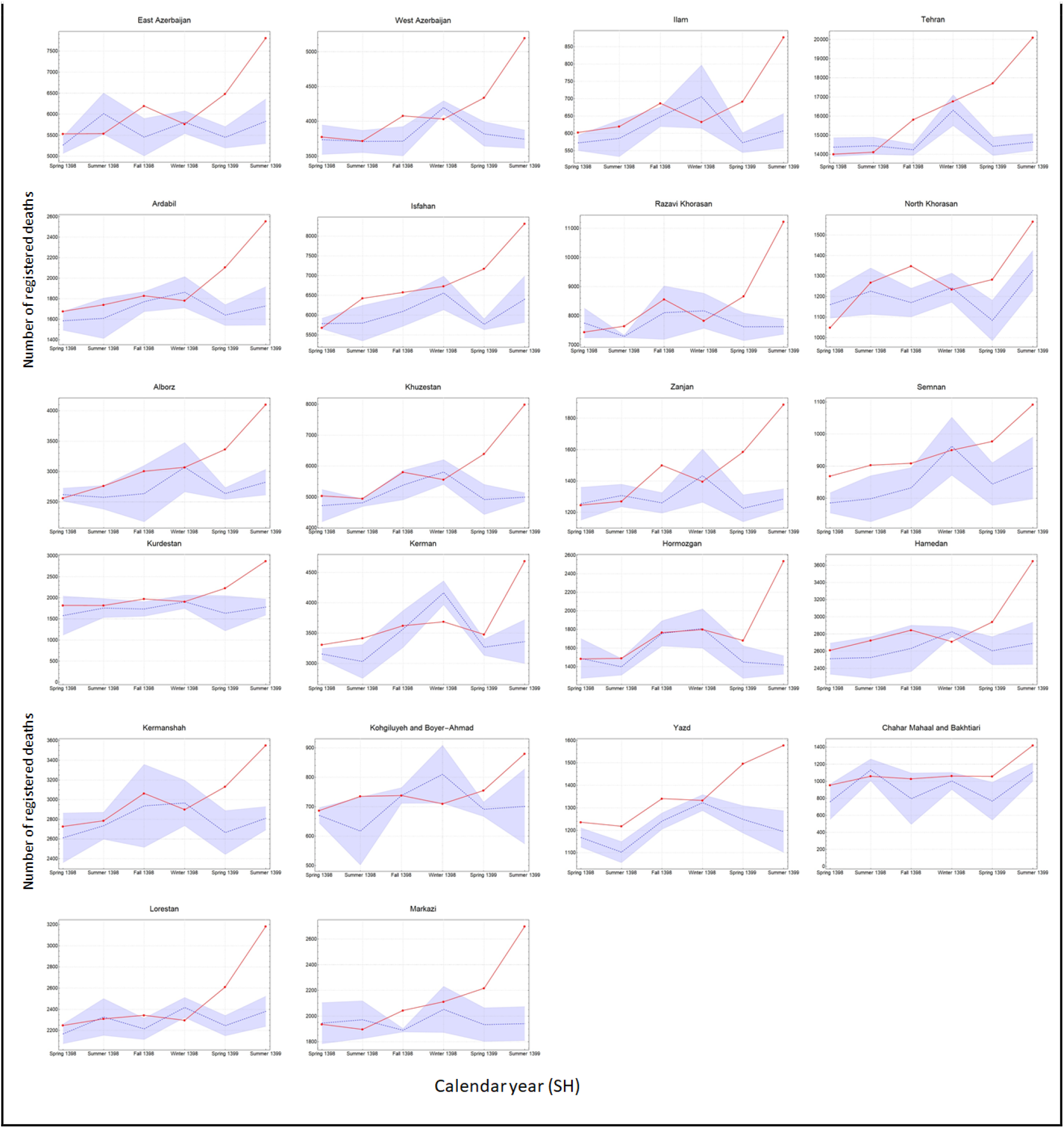
The pattern of excess mortality from spring 1398 SH (21 March 2019) to summer 1399 SH (21 September 2020) in **(a)** 5 provinces that experienced significant levels of excess mortality from winter 1398 SH to summer 1399 SH, **(b)** 4 provinces that experienced high mortality rates in either spring or summer 1399 SH, and **(c)** the 22 remaining provinces that experienced a growing excess mortality trend from spring 1399 SH to summer 1399 SH.

### Estimating the level of exposure across the population

Assuming that all the excess deaths since winter 1398 SH onward are directly associated with the COVID-19 epidemic, we applied a population-weighted Infection Fatality Ratio (IFR) [14] to estimate the level of exposure in each province (see **Methods** section) by the end of summer 1399 SH (see **Table 2**). We find that 13 (out of 31) provinces had nearly 30% exposure or above with Qom and Golestan having the highest percentage of individuals exposed at 57% (95%CI: 44% - 69%) and 56% (95%CI: 44% - 69%), respectively (see **Figure 4**).

**Table 2:** Excess mortality associated with COVID-19 and estimated level of exposure in each province. We use the age-structure of each province from the Statistical Center of Iran [29] to calculate their corresponding population-weighted IFR. Numbers in (.) show the 95% confidence intervals.

| Provinces | Excess deaths | %IFR | Number of individuals exposed | %Exposed |
| --- | --- | --- | --- | --- |
| East Azerbaijan | 3.0K (2.2K - 3.8K) | 0.30 (0.28 - 0.32) | 1.01M (0.72M - 1.29M) | 25 (18 - 32) |
| West Azerbaijan | 2.0K (1.7K - 2.3K) | 0.24 (0.22 - 0.25) | 0.83M (0.64M - 1.03M) | 24 (18 - 30) |
| Ardabil | 1.3K (1.0K - 1.6K) | 0.26 (0.25 - 0.28) | 0.49M (0.36M - 0.61M) | 37 (28 - 47) |
| Isfahan | 3.3K (2.6K - 4.0K) | 0.29 (0.27 - 0.31) | 1.13M (0.81M - 1.45M) | 21 (15 - 27) |
| Alborz | 2.0K (1.7K - 2.3K) | 0.24 (0.22 - 0.25) | 0.84M (0.64M - 1.05M) | 29 (22 - 36) |
| Ilam | 0.4K (0.3K - 0.5K) | 0.23 (0.22 - 0.25) | 0.17M (0.13M - 0.21M) | 28 (21 - 34) |
| Bushehr | 0.7K (0.6K - 0.7K) | 0.18 (0.17 - 0.19) | 0.38M (0.30M - 0.45M) | 30 (24 - 36) |
| Tehran | 8.8K (7.8K - 9.7K) | 0.28 (0.26 - 0.30) | 3.09M (2.24M - 3.94M) | 22 (16 - 28) |
| South Khorasan | 0.1K (0.0K - 0.1K) | 0.26 (0.24 - 0.29) | 0.03M (0.02M - 0.04M) | 4 (3 - 5) |
| Razavi Khorasan | 4.6K (3.9K - 5.4K) | 0.23 (0.22 - 0.25) | 2.00M (1.53M - 2.47M) | 29 (22 - 36) |
| North Khorasan | 0.4K (0.2K - 0.6K) | 0.22 (0.21 - 0.24) | 0.19M (0.15M - 0.24M) | 22 (17 - 27) |
| Khuzestan | 4.5K (3.9K - 5.1K) | 0.19 (0.28 - 0.20) | 2.33M (1.85M - 2.81M) | 47 (37 - 57) |
| Zanjan | 1.0K (0.8K - 1.1K) | 0.28 (0.26 - 0.30) | 0.35M (0.26M - 0.44M) | 32 (23 - 40) |
| Semnan | 0.3K (0.2K - 0.5K) | 0.27 (0.25 - 0.29) | 0.12M (0.09M - 0.16M) | 16 (12 - 21) |
| Sistan and Baluchistan | 0.9K (0.6K - 1.2K) | 0.13 (0.12 - 0.13) | 0.73M (0.62M - 0.84M) | 24 (20 - 27) |
| Fars | 1.8K (1.5K - 2.2K) | 0.26 (0.24 - 0.28) | 0.70M (0.52M - 0.88M) | 14 (10 - 17) |
| Qazvin | 1.6K (1.1K - 2.1K) | 0.25 (0.23 - 0.27) | 0.64M (0.48M - 0.80M) | 48 (36 - 60) |
| Qom | 1.6K (1.4K - 1.8K) | 0.21 (0.19 - 0.22) | 0.79M (0.62M - 0.96M) | 57 (44 - 69) |
| Kurdistan | 1.7K (1.1K - 2.3K) | 0.25 (0.23 - 0.26) | 0.68M (0.51M - 0.85M) | 41 (31 - 51) |
| Kerman | 1.5K (1.0K - 2.0K) | 0.22 (0.20 - 0.23) | 0.72M (0.56M - 0.87M) | 21 (13 - 26) |
| Kermanshah | 1.2K (0.9K - 1.5K) | 0.27 (0.25 - 0.29) | 0.45M (0.33M - 0.56M) | 22 (16 - 28) |
| Kohgiluyeh and Boyer-Ahmad | 0.2K (0.1K - 0.4K) | 0.20 (0.19 - 0.22) | 0.12M (0.09M - 0.15M) | 16 (12 - 19) |
| Lorestan | 1.2K (0.9K - 1.4K) | 0.25 (0.23 - 0.27) | 0.47M (0.35M - 0.58M) | 26 (20 - 32) |
| Mazandaran | 3.7K (3.3K - 4.1K) | 0.31 (0.29 - 0.34) | 1.18M (0.83M - 1.53M) | 35 (25 - 45) |
| Markazi | 1.0K (0.8K - 1.3K) | 0.32 (0.29 - 0.34) | 0.33M (0.23M - 0.42M) | 22 (16 - 29) |
| Hormozgan | 1.4K (1.1K - 1.6K) | 0.17 (0.16 - 0.18) | 0.81M (0.66M - 0.96M) | 42 (34 - 49) |
| Hamedan | 1.3K (0.9K - 1.7K) | 0.30 (0.28 - 0.32) | 0.43M (0.31M - 0.55M) | 24 (17 - 31) |
| Yazd | 0.6K (0.5K - 0.8K) | 0.24 (0.22 - 0.26) | 0.27M (0.20M - 0.33M) | 21 (16 - 27) |
| Chaharmahal and Bakhtiari | 0.6K (0.3K - 0.9K) | 0.24 (0.22 - 0.26) | 0.25M (0.19M - 0.31M) | 26 (19 - 32) |
| Golestan | 2.3K (1.9K - 2.6K) | 0.20 (0.19 - 0.22) | 1.12M (0.87M - 1.36M) | 56 (44 - 69) |
| Gilan (Guilan) | 3.1K (2.5K - 3.8K) | 0.36 (0.34 - 0.39) | 0.86M (0.57M - 1.14M) | 33 (22 - 44) |
| Nationwide | 58.9K (46.9K - 69.5K) | 0.25 (0.24 - 0.27) | 23.0M (17.2M - 28.7M) | 27 (20 - 34) |

**Figure 4:**
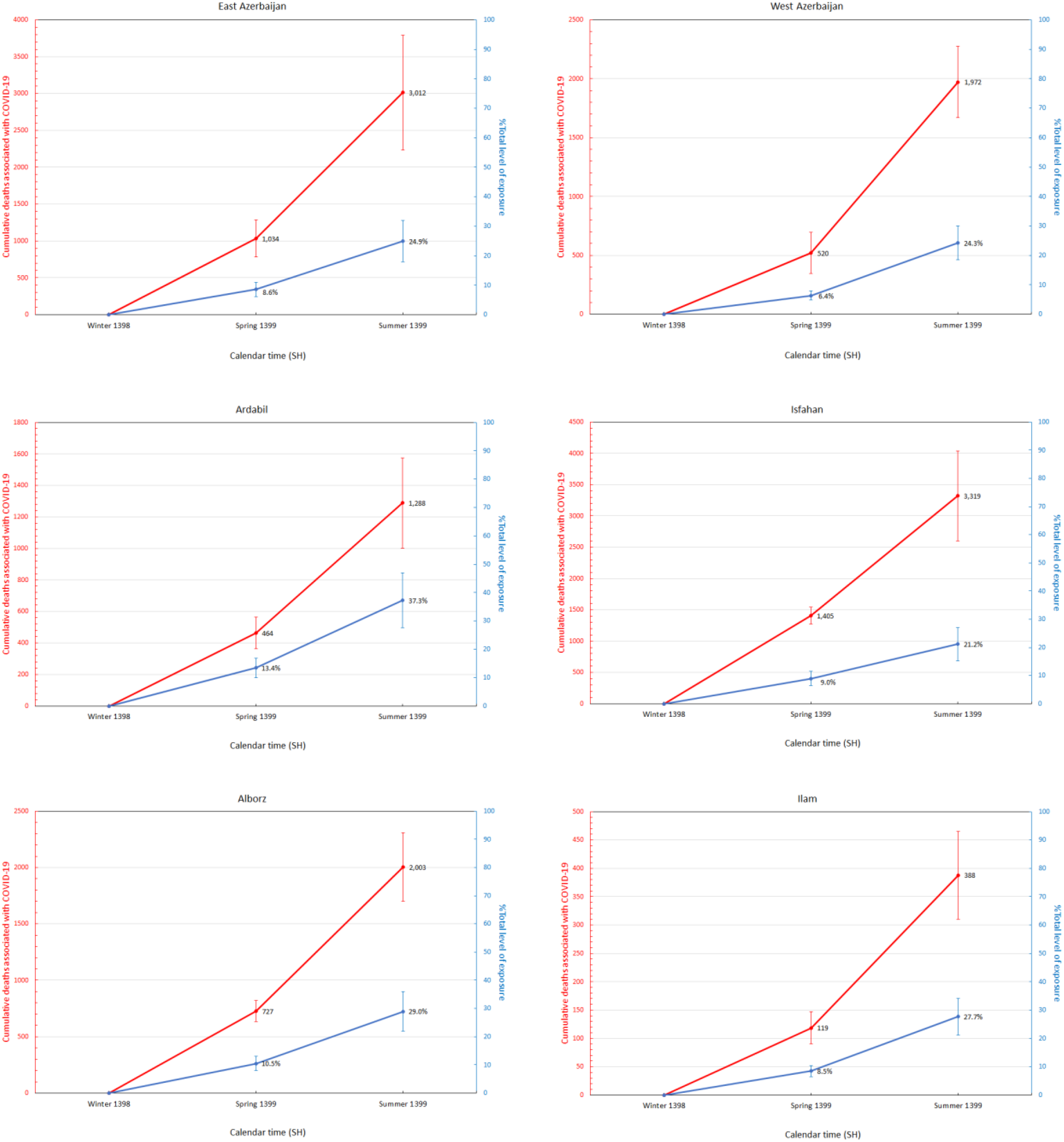
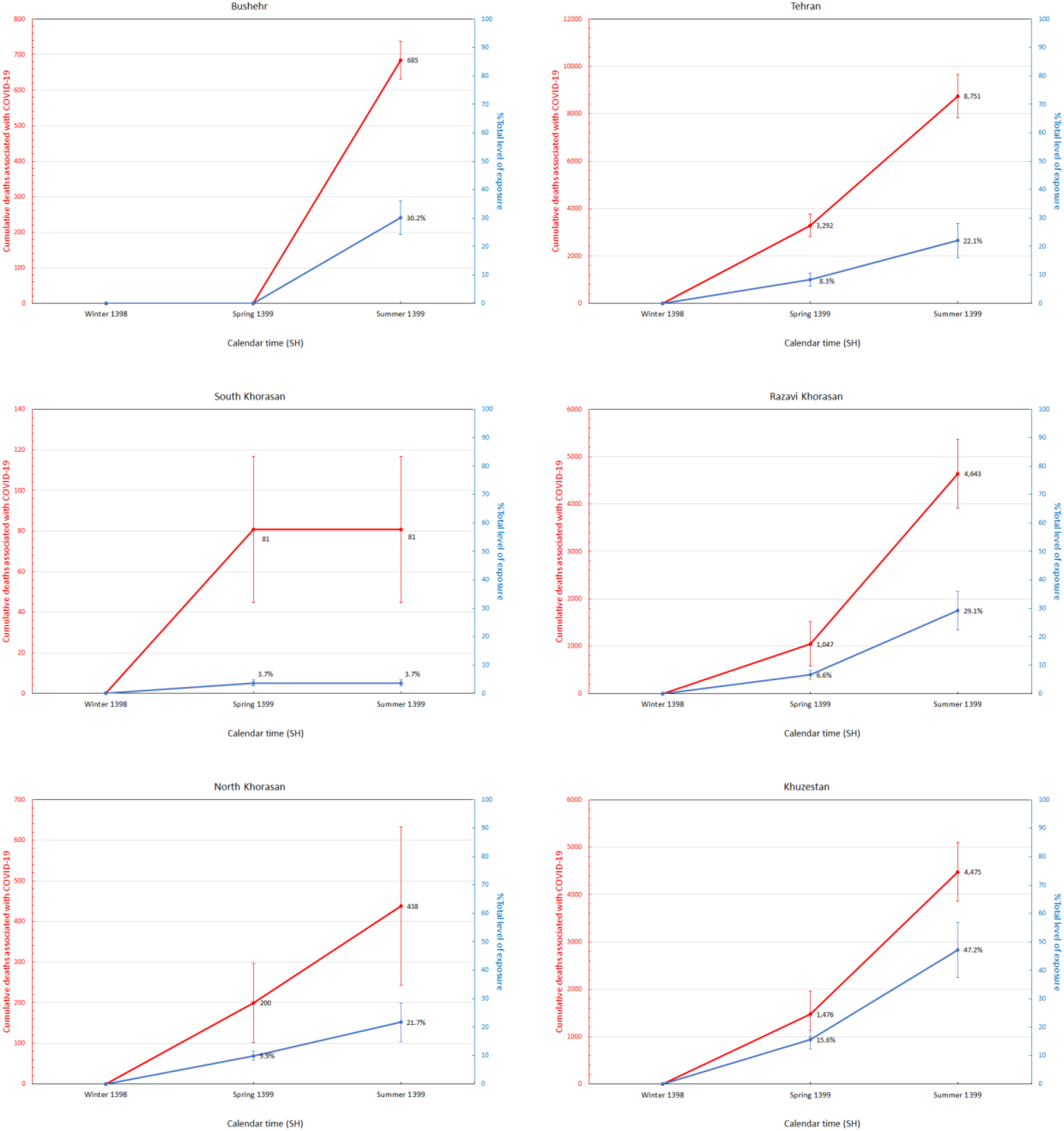
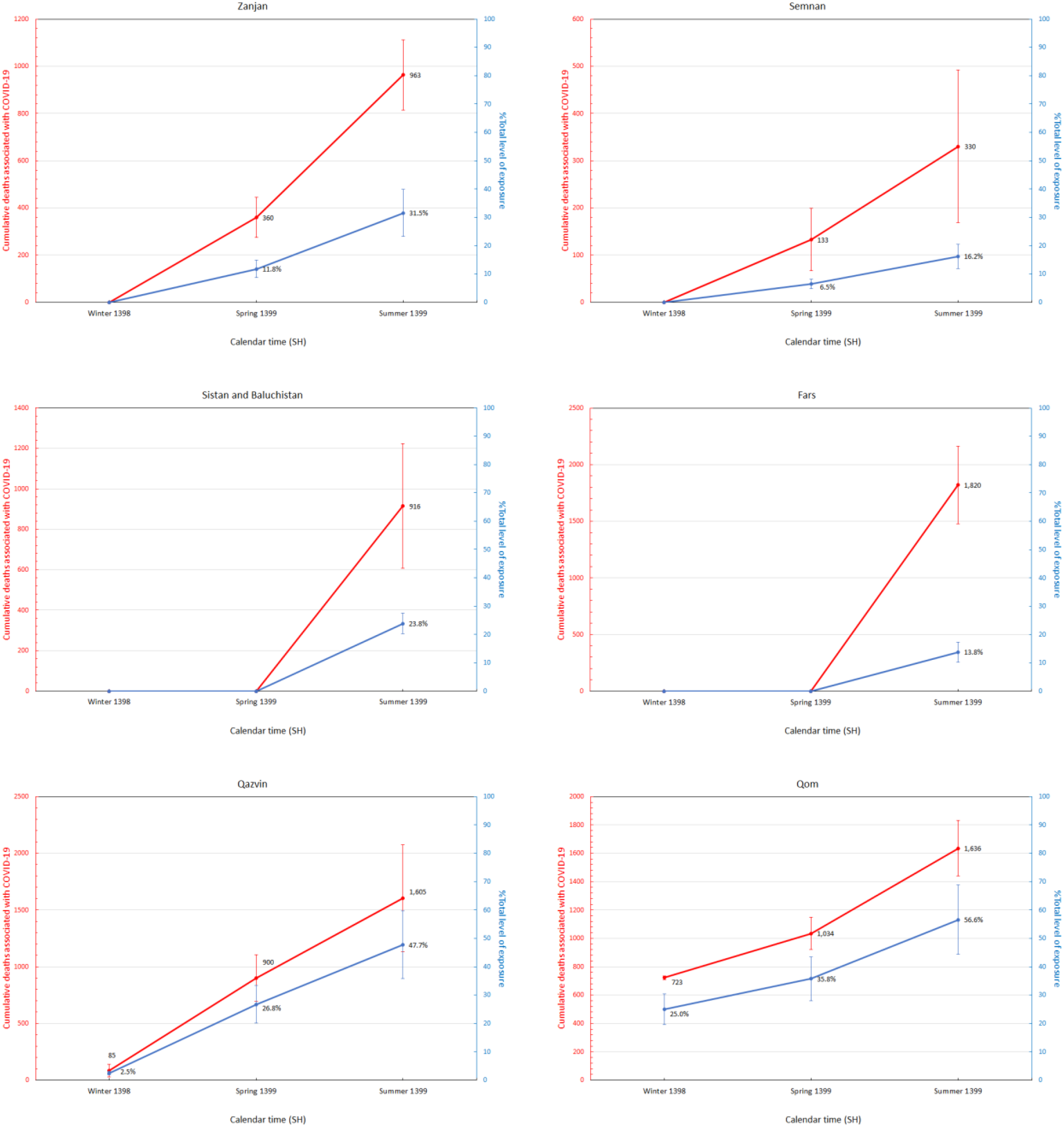
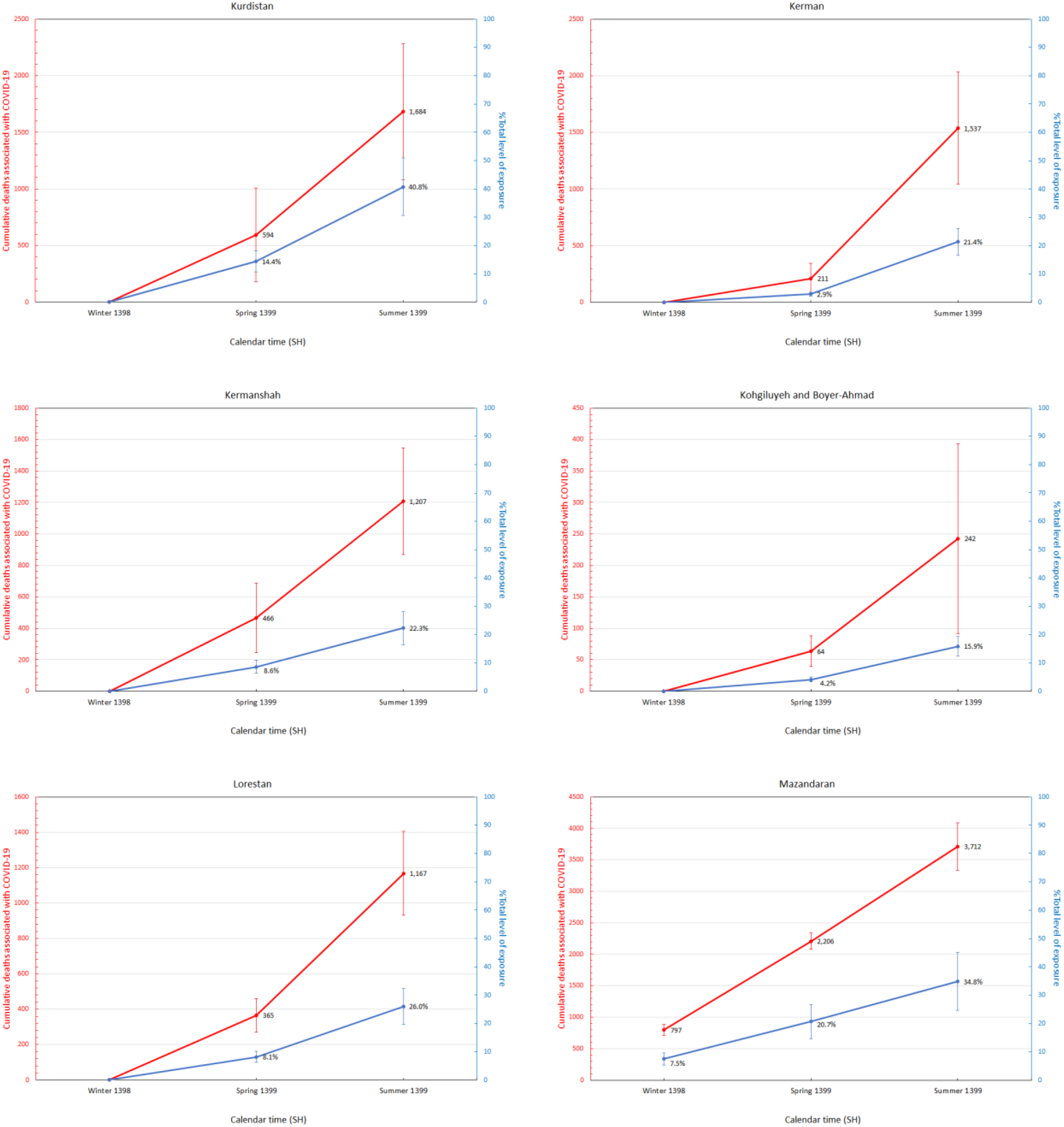
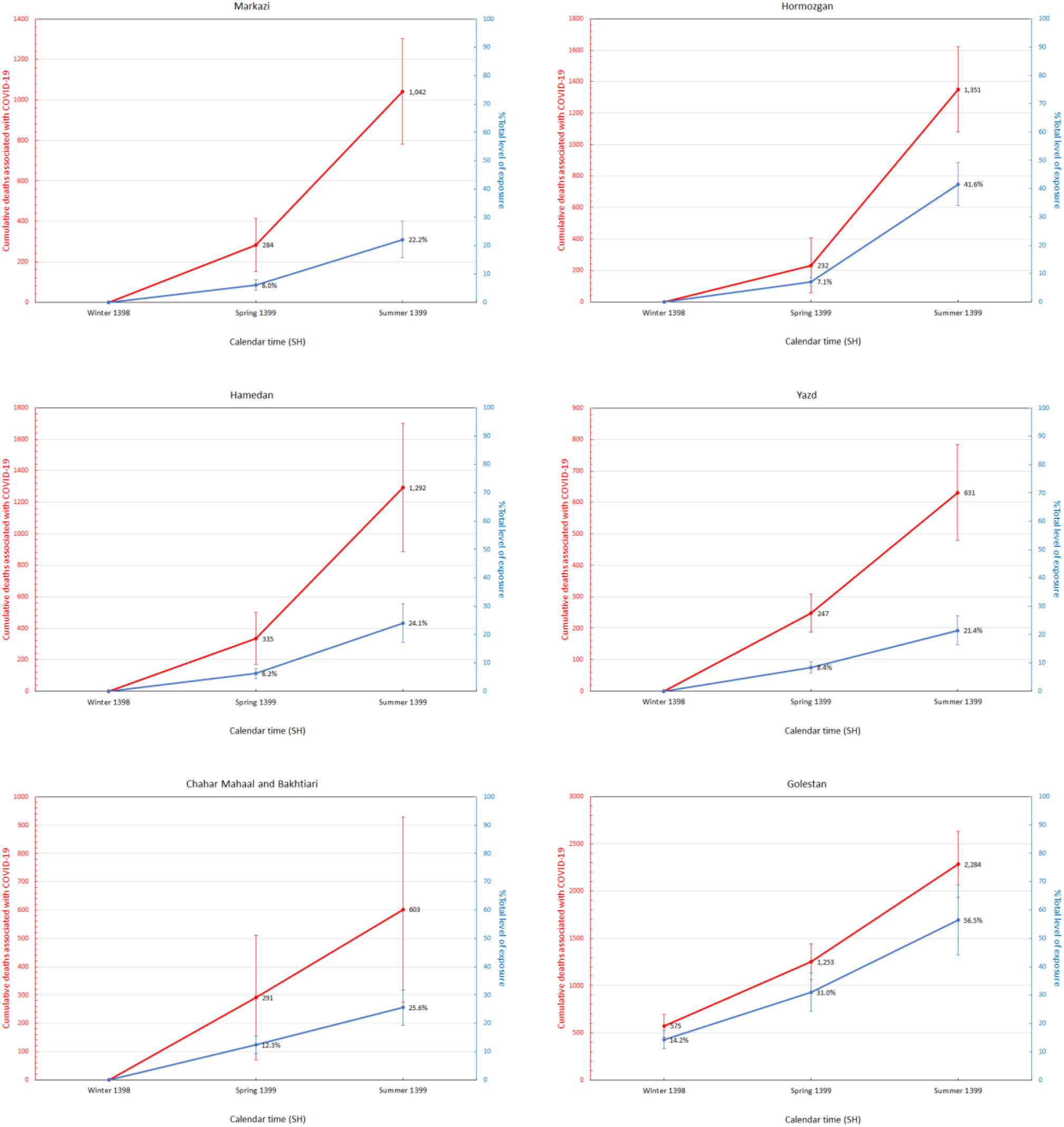
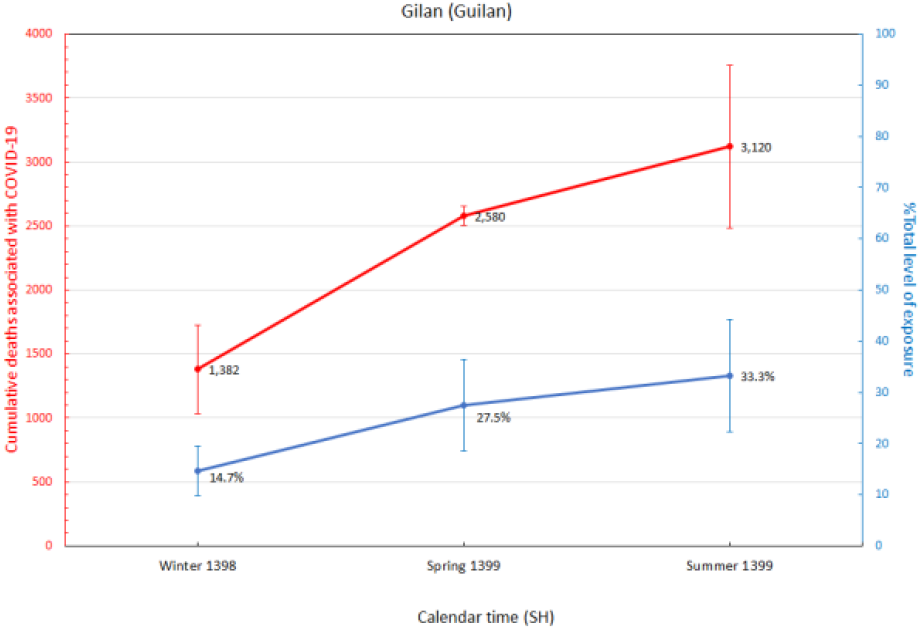
Cumulative number of COVID-19-related deaths (red) and total fraction of individuals exposed to SARS-CoV-2 in each province from winter 1398 SH to summer 1399 SH.

By combining the estimated number of deaths associated with COVID-19 based on excess mortality and the number of individuals exposed to SARS-CoV-2 in each province, we can reconstruct the nationwide trend of seasonal COVID-19-associated infections and deaths (see **Figure 5**). We find that the corrected nationwide COVID-19-related deaths during each season is approximately 2.5 times higher than the number of confirmed deaths from MoHME and that both estimates follow a similar trajectory over time with particularly elevated levels of excess mortality during winter (i.e. approximately 2.8 times higher than the confirmed deaths at the time) which may indicate that a larger portion of deaths during the early stages of the epidemic were under-counted as also noted in our previous analysis [13].

**Figure 5:**
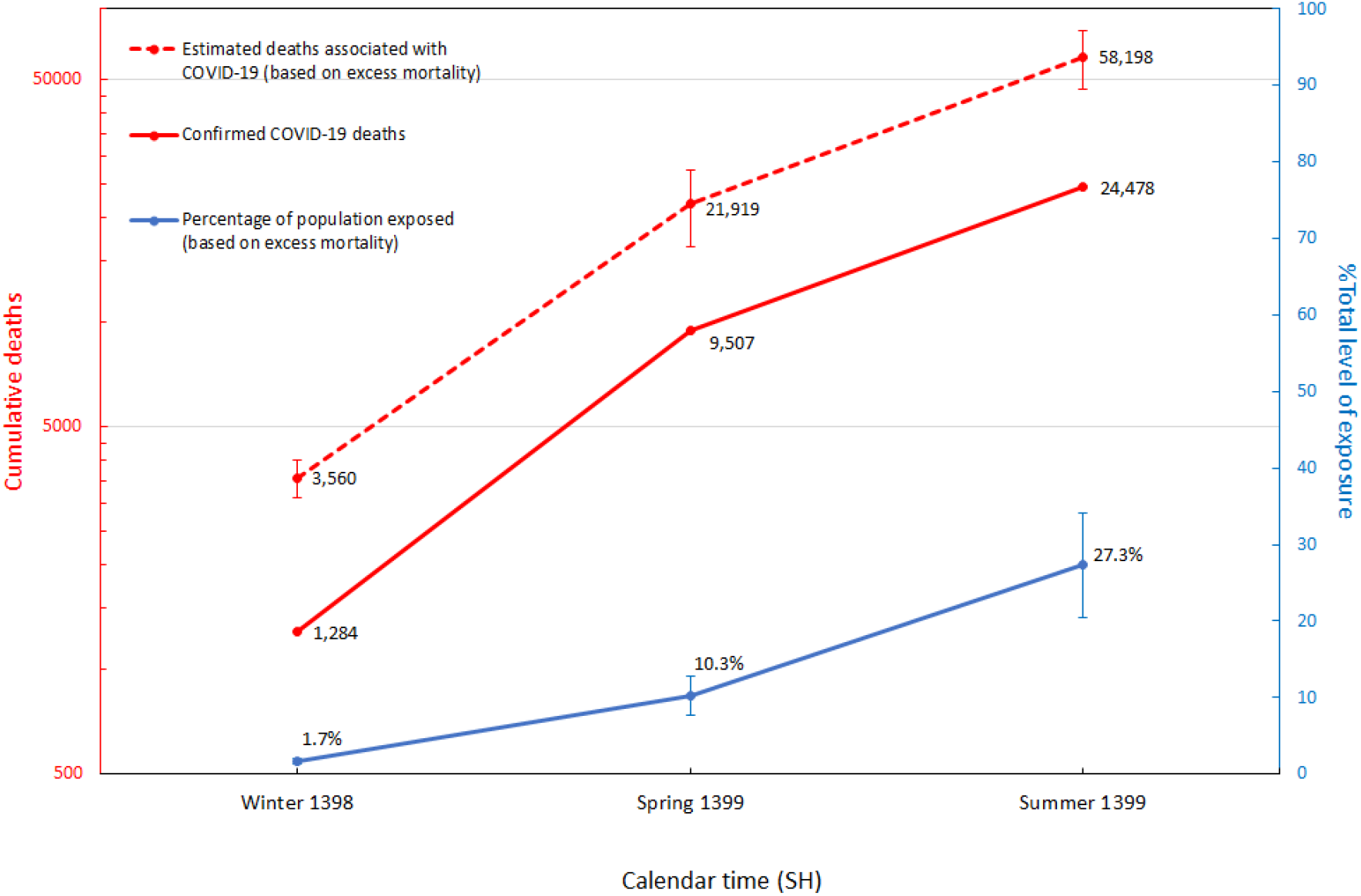
Estimated number of deaths and exposures to COVID-19 based on excess mortality data from winter 1398 SH to summer 1399 SH. Confirmed COVID-19 deaths are collected from the daily reports by MoHME.

## Discussion

In this work, we compared mortality rates in 31 provinces of Iran from winter 1398 SH to summer 1399 SH with the aim to detect and measure excess deaths related to the COVID-19 epidemic. Our findings suggest a total of 58.9 (95%CI: 46.9 - 69.5) thousand COVID-19-related deaths and up to 23.0 (95%CI: 17.2 - 28.7) million individuals exposed to SARS-CoV-2 by 21 September 2020 with some of the hardest-hit provinces such as Qom and Golestan reaching up to approximately 57% (95%CI: 44% - 69%) exposure levels.

We are planning to continue monitoring the all-cause mortality data in the coming seasons as this is currently the only detailed provincial data available to gauge the level of spread across the country and assess possible under-counting of COVID-19-related deaths. Given that the epidemic was never fully brought under control with three major peaks in late-March, mid-July, and late-November that overwhelmed hospitals (as of 30 November 2020), some provinces are experiencing among the highest known population-level prevalence in the World, comparable to Manaus, Brazil [15,16].

Recent reports from the Iranian health officials indicate that up to half of suspect hospitalised cases may not be tested due to shortage of test kits but are being treated as COVID-19 patients based on their clinical symptoms. This highlights the large degree of under-reporting in the actual number of COVID-19 cases and deaths which also seems to be aligned with our findings that the actual number of COVID-19-related deaths could be up to around 2.5 times higher than the number of confirmed deaths. While COVID-19 is directly responsible for the majority of excess deaths during the epidemic, it is possible that not all excess deaths are directly attributable to infection from the disease [17]. We also note that NOCR’s registration system might be strained when facing a potentially massive backlog of uncounted death certificates and prone to temporary under-counting. Our earlier analysis shows that deaths due to other causes such as traffic injuries have only dropped by a few hundred compared to previous years (data can be obtained from the Iranian Legal Medicine Organization [18]) and is not a major contributor to the changing trend in excess mortality during the epidemic [19].

None of the nationwide serology reports have been published except for the earliest report in late-spring [20] which estimated that around 10-15 million Iranians (11.9%-17.9% of the population) have been infected by late spring. Later in mid-summer, a statement from MoHME’s deputy minister of research and technology was released by the press [21] that approximately 25 million (29.8% of the population) have so far been exposed to the virus based on their recent seroepidemiological analysis. In our earlier analysis [13], we reconstructed the early transmission dynamics of the Iranian epidemic and projected that as the second peak emerges in summer there will be a total of 9 million individuals (10.7% of the population) recovered by 14 July 2020 which is very close to our current 10.3% estimates for nationwide exposure levels based on excess mortality data by 20 June 2020. Although we cannot independently verify the validity of any of the nationwide seroprevalence studies released to the press, they seem to be in general agreement with our estimates based on seasonal excess mortality data during winter and spring. Assuming that the ratio between excess mortality and confirmed COVID-19-related deaths remains the same over time (i.e. approximately 2.5), by 25 October 2020 there would have been a total of approximately 84 thousand COVID-19-related deaths (based on the 32,616 confirmed deaths at the time) and 33 million Iranians exposed to SARS-CoV-2 which is very close to the 35 million estimate based on the nationwide seroprevalence studies at the time [22].

The only systematic seroprevalence study in Iran took place in Gilan during April 2020 with an estimated prevalence of 21.0% (95%CI: 17.6% - 24.7%) across 528 household and community samples corresponding to an IFR of 0.09% (95%CI: 0.08% - 0.12%) [23]. More recent systematic reviews and meta-analysis of global seroprevalence of SARS-CoV-2 suggests that this seroprevalence study has moderate levels of bias and has overestimated the true prevalence by approximately a factor of 3.9 [24,25]. This implies that the corrected IFR of Gilan is more likely around 0.35% which is much closer to our 0.36% (95%CI: 0.34% - 0.39%) population-weighted IFR estimate. We found that the excess mortality was up to 2.6 times higher during the period close to when the seroprevalence study was carried out. Thus, we conclude that the true prevalence in Gilan by the end of winter 1398 SH was 14.7% (95%CI: 9.9% - 19.5%).

We also demonstrated that the mean excess mortality rate across the country has increased by more than tenfold since the start of the epidemic and that all provinces had high mortality rates for at least one season as a consequence of the COVID-19 epidemic. In particular, most of the provinces that had a significant number of excess deaths in more than one season, experienced a surge in the death toll following the initial surge back in winter and/or spring which indicates that the epidemic was never fully brought under control. Given the record-high number of confirmed COVID-19-related deaths during fall 2020, we predict that the excess mortality counts will continue to increase across the country once NOCR updates the data for fall 1399 SH.

We previously showed, through an epidemiological and phylogenetic analysis on individuals with a travel link to Iran, that there were significant levels of under-reporting of cases and deaths, particularly at the early stages of the epidemic which started back in mid-to late-January [11]. By comparing the mortality trends in provinces with significant number of recorded deaths both in fall (13 out of 31 provinces) and winter (5 out of 31 provinces) 2019-20, we determined that the unusually high mortality rates from 23 September to 21 December 2019 were not linked to any potential cryptic transmission of the virus across the country. More detailed investigation into the causes of deaths and time-resolved monitoring for acute respiratory infections are required to determine the key source(s) of the spike during this period.

We note that to estimate the mean IFR for each province, we re-weighted the IFR of each age group by its respective share of the total population in each province. This procedure relies on the assumption that contagion was homogeneous across all age groups which may not be true due to reasons such as the heterogeneity in the burden of the epidemic in nursing homes [14] and potential age-varying susceptibility to infection by SARS-CoV-2 [26]. However, a recent analysis shows that the increase in mortality by age from COVID-19 strongly resembles the age pattern of all-cause mortality [27]. Also, our provincial IFR estimates ranging from lowest, 0.13%, in Sistan and Baluchistan to highest, 0.36%, in Gilan are well-aligned with those of many other low-income countries [28].

## Methods

To calculate the expected number of deaths in one season of a particular year for one province we apply a linear regression model using the reported deaths from that particular season starting from the year 1394 SH until one year prior to the year of interest and consider anything above 2 standard deviations units from the mean (95% confidence interval) to be a significant level of excess mortality. Given the limited number of data points per year, i.e. one data point for every season, we avoid using more complex models such as the generalised linear models which may demand more parameters to fit the data. Nevertheless, using a standard 5-year average to calculate the expected number of deaths may also bias our estimates as it does not correctly account for significant changes in the excess mortality trend over time due to, for instance, a changing population size or other secular trends that are present in NOCR’s data. We also note that although some provinces show a seasonal trend in mortality with higher death rates during fall and winter compared to spring and summer, such patterns are not readily detectable for every province (i.e. variations during each season are high). Therefore, we also avoid using a periodic regression model as it may bias our estimates for expected seasonal deaths [12].

Assuming an equal attack rate across all age groups, we use the province-specific age demographic (data collected from the Statistical Center of Iran [29]) to calculate the corresponding population-weighted IFR [14]. To find the number of individuals exposed to the virus in each province, *Z*, we took our estimated number of COVID-19-associated deaths based on excess mortality, *X*, and divide that by the IFR, *Y*, such that the corresponding variance (Var) in *Z* is given by

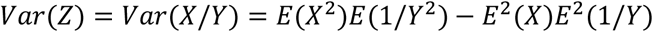

where *X* is a normally distributed number, and the standard deviation of *Y* is computed using the Delta method (see below). Thus, the inverse of *Y* is taken as a random number from the reciprocal normal distribution.

To calculate the standard deviation in *Y*, we took the age-stratified 95%CI estimates from [14] to find the age-specific IFR, i.e. *IFR*_*i*_, corresponding to age-group *i*, and calculate the standard deviation, *σ*_*i*_, using the Delta method, i.e. *σ*_*i*_ *=* (95% *upper limit* − 95% *lower limit*) / *z*_1−*α*/2_ where *z*_1−*α*/2_is the (1 − *α*/2) quantile of the standard normal distribution. Then, assuming that deaths occur homogeneously across all age groups, the population-weighted IFR of each province with a total population size, *P*, is given by 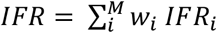 with a standard deviation 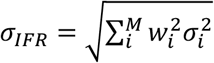 such that *M* is the total number of age-groups and each group, *i*, has a population size *p*_*i*_ and *w*_*i*_ *= p*_*i*_/*P*.

## Data Availability

All the datasets used in this study are publicly available at the National Organization for Civil Registration (NOCR) and can be found on sabteahval.ir website.

## Acknowledgement

MG would like to thank Kaveh Madani for his constructive comments on the analysis of excess mortality. MG and AK designed the analysis. MG wrote the manuscript. MG and AKad carried out the analysis. All authors reviewed and edited the manuscript.

